# Community structure mediates Sabin 2 polio vaccine virus transmission

**DOI:** 10.1101/2020.07.01.20144501

**Authors:** M Famulare, W Wong, R Haque, JA Platts-Mills, P Saha, AB Aziz, T Ahmed, MO Islam, MJ Uddin, AS Bandyopadhyay, M Yunus, K Zaman, M Taniuchi

## Abstract

Since the global withdrawal of Sabin 2 oral poliovirus vaccine (OPV) from routine immunization, the Global Polio Eradication Initiative (GPEI) has reported multiple circulating vaccine-derived poliovirus type 2 (cVDPV2) outbreaks. cVDPV2 outbreaks are controlled with mass Sabin 2 OPV vaccination campaigns, which carry a small but serious risk of seeding future cVDPV2 outbreaks. Accurate forecasting models are essential for quantifying current and future cVDPV2 outbreak risk following Sabin 2 OPV campaigns, but it is unclear whether household community structure influences vaccine virus transmission or is relevant for assessing cVDPV2 outbreak risk. Here, we developed an agent-based model of Sabin 2 vaccine transmission to assess the role of household community structure on vaccine virus transmission following a mass OPV campaign performed in Matlab, Bangladesh. Household community structure strongly limits vaccine virus transmission to local community members and ignoring it overestimates outbreak risk in terms of emergence probability, duration, and epidemic size.

## Introduction

The Global Polio Eradication Initiative (GPEI) is one of the largest public health interventions dedicated to the eradication of a specific disease (1, 2). Since its launch in 1988, the global incidence of poliomyelitis has fallen by more than 99% and wild poliovirus (WPV) types 2 and 3 have been eradicated (3, 4). The success of the GPEI is largely due to the effectiveness of the live-attenuated oral poliovirus vaccines (OPV) for preventing disease and transmission and its deployment in mass campaigns (5). However, continued Sabin 2 OPV-related outbreaks of circulating vaccine-derived poliovirus type 2 (cVDPV2) since the withdrawal of Sabin 2 OPV from routine immunization schedules are of major concern (6). cVDPV2 outbreaks are facilitated by persistently low population-level immunity. These outbreaks are controlled through mass vaccination with Sabin 2 OPV, which bolsters immunity but can seed future cVDPV2 outbreaks (7–9).

As global immunity against Sabin 2 declines, controlling and preventing future outbreaks will become more difficult with the existing vaccine options. Mathematical models are useful tools for assessing transmission and outbreak risk but a major challenge is the incorporation of biological realism regarding population structure, immunity, transmissibility, and contact structure (10–13). Dynamic, agent-based models can be designed to simulate the complex interplay between these factors and have been deployed to characterize poliovirus transmission across different populations (14, 15). However, there is little consensus on the importance of social contact structure for understanding poliovirus vaccine transmission. Models typically simulate transmission using the laws of mass action, which assumes all individuals have the same exposure risk and contact probability. This assumption is particularly prevalent in disease forecasting models (16–18). However, theoretically-motivated poliovirus models show social contact structure has the potential to affect transmission and alter model predictions (19), but whether such concerns are relevant for assessing empirical vaccine transmission or evaluating future cVDPV2 outbreak risk is uncertain.

Clinical studies with OPV present a unique opportunity to assess the role of social contact structure based on empirical vaccine transmission. We previously conducted a clinical trial in Matlab, Bangladesh to monitor community transmission and assess changes in Sabin 2 transmission following reintroduction of Sabin 2 from a mass monovalent Sabin 2 OPV (mOPV2) vaccination campaign (20). Matlab is a highly immunized setting where individuals are organized into strongly defined household and community structures. By utilizing the data collected during the vaccine trial and information regarding the demographic structure of Matlab, we developed and calibrated a mathematical model to assess whether the constraints on social contact structure imposed by household and community structure alters poliovirus vaccine transmission. Understanding this is critical for our understanding of poliovirus transmission and for forecasting future cVDPV2 outbreak and transmission risk.

## Results

### Brief clinical trial and model overview

We developed an agent-based Sabin 2 transmission model (**Fig. 1**) that simulates vaccine transmission following a monovalent Sabin 2 OPV (mOPV2) clinical trial performed in Matlab, Bangladesh (20) (**Fig. S1**). The clinical trial involved 67 villages and was split into two phases: 1) a nine-month enrollment and routine immunization with tOPV (trivalent OPV, contains Sabin 1, 2, and 3), bOPV (bivalent OPV, Sabin 1 and 3) and one dose of inactivated poliovirus vaccine (IPV), or bOPV + two IPV doses, and 2) a 22 week surveillance period following an mOPV2 vaccination challenge campaign targeting infants, the two youngest household contacts of said infant, and community members under five years old.

**Fig. 1.**
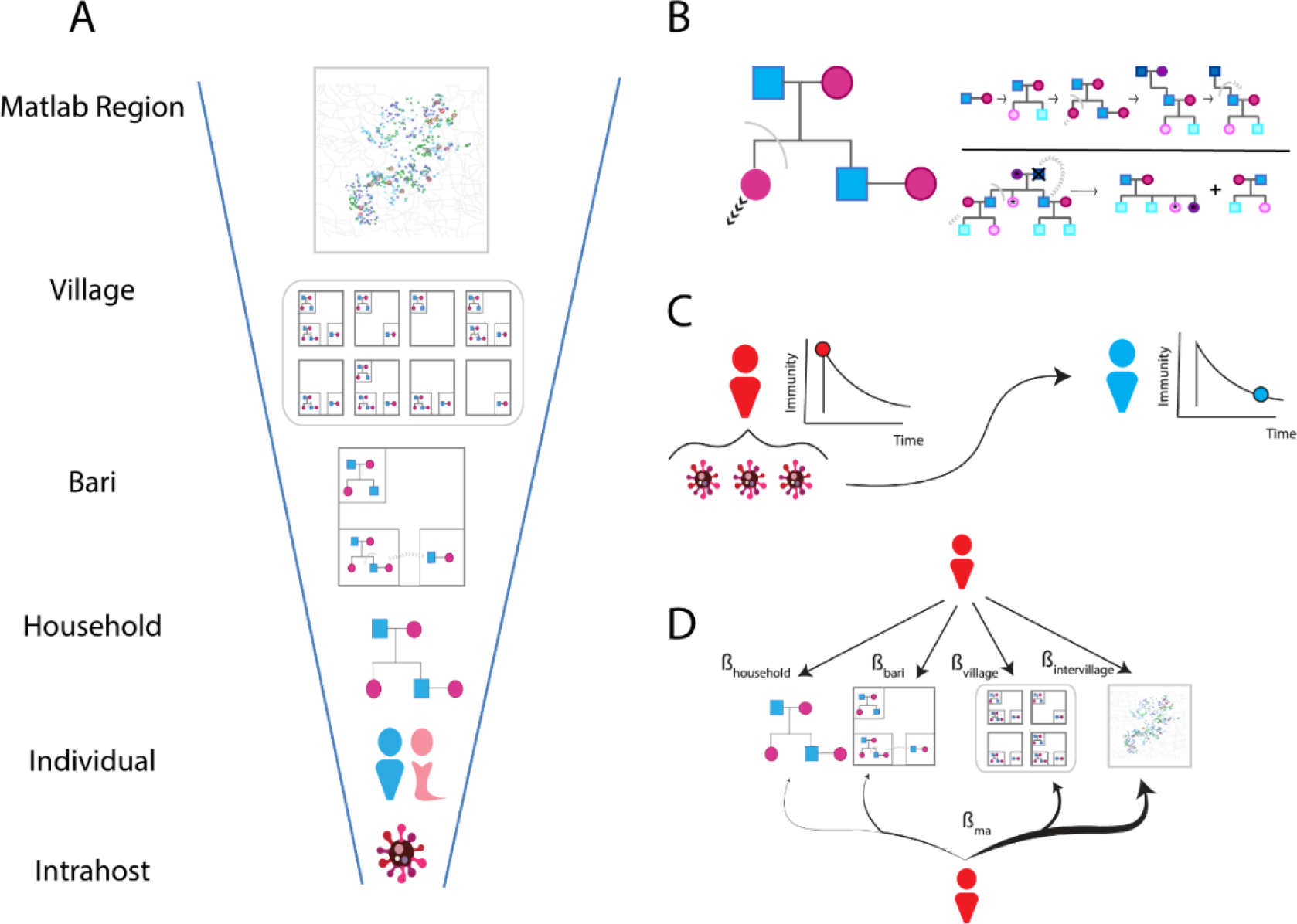
Model Structure. **A**) Simulated populations are organized into a series of hierarchical, multiscale demographic structures that define the Matlab Region. The *bari* is an intergenerational living arrangement of related household members specific to rural Bangladesh. **B**) Household demographic model. Individuals are grouped into households that whose composition changes as individuals are born, marry, and leave (*top right*). The death of the household head triggers secession and splits the household along familial lineages (*bottom right*). **C**) Dose-response infection model. Successful transmission depends on the viral exposure dose per contact and the immunity of the contacted individual. Infected individuals receive a boost in immunity that wanes over time. Infected individuals shed and transmit viruses to other individuals through contact. The total exposure dose depends on the viral shedding concentration and the average fecal-oral dose per contact. **D**) Transmission models. Contacts are allocated using mass action (*bottom*) or multiscale transmission (*top*). Mass action uses a single contact rate β_ma_ to distribute contacts across demographic scales proportional to their size. The multiscale transmission model utilizes demographic structure by allocating transmission with four scale-specific contact rates (β_household_, β_bari,_ β_village_, β_intervillage_).

Demographic structure was simulated using a household demographic model that groups individuals into households that evolve using a set of anthropological rules governing household formation and secession (**Methods**). This model was used to replicate the household size and household contact ages reported in the clinical trial (20) and the Bangladeshi age pyramid (**Fig. S2**). All simulations contained an average of 83,000 individuals distributed throughout 45 villages with the same size distribution as the bOPV-assigned villages in the clinical trial.

### Fitting contact structure to clinical trial data

Model fits were first made using two different contact structures: 1) mass action, where infectious contacts are made randomly throughout the population, and 2) multiscale, where infectious contacts are made at different rates to each demographic scale (household, bari, village, region). We calibrated the multiscale and mass action frameworks to the data collected during the routine immunization/enrollment phase and the post-mOPV2 surveillance period. When calibrated to both data sets, mass action failed to replicate the clinical trial data (**Fig. 2, Fig. S3**). Under this fully-calibrated mass action model, transmission was dominated by intervillage transmission events; less than 1% of all transmissions occurred at the household, bari, or village sublevels. The fully-calibrated mass action model predicted no shedding in any of the longitudinal cohort-shedding profiles of non-mOPV2 challenged individuals (**Fig. 2B-C**, *green*).

**Fig. 2.**
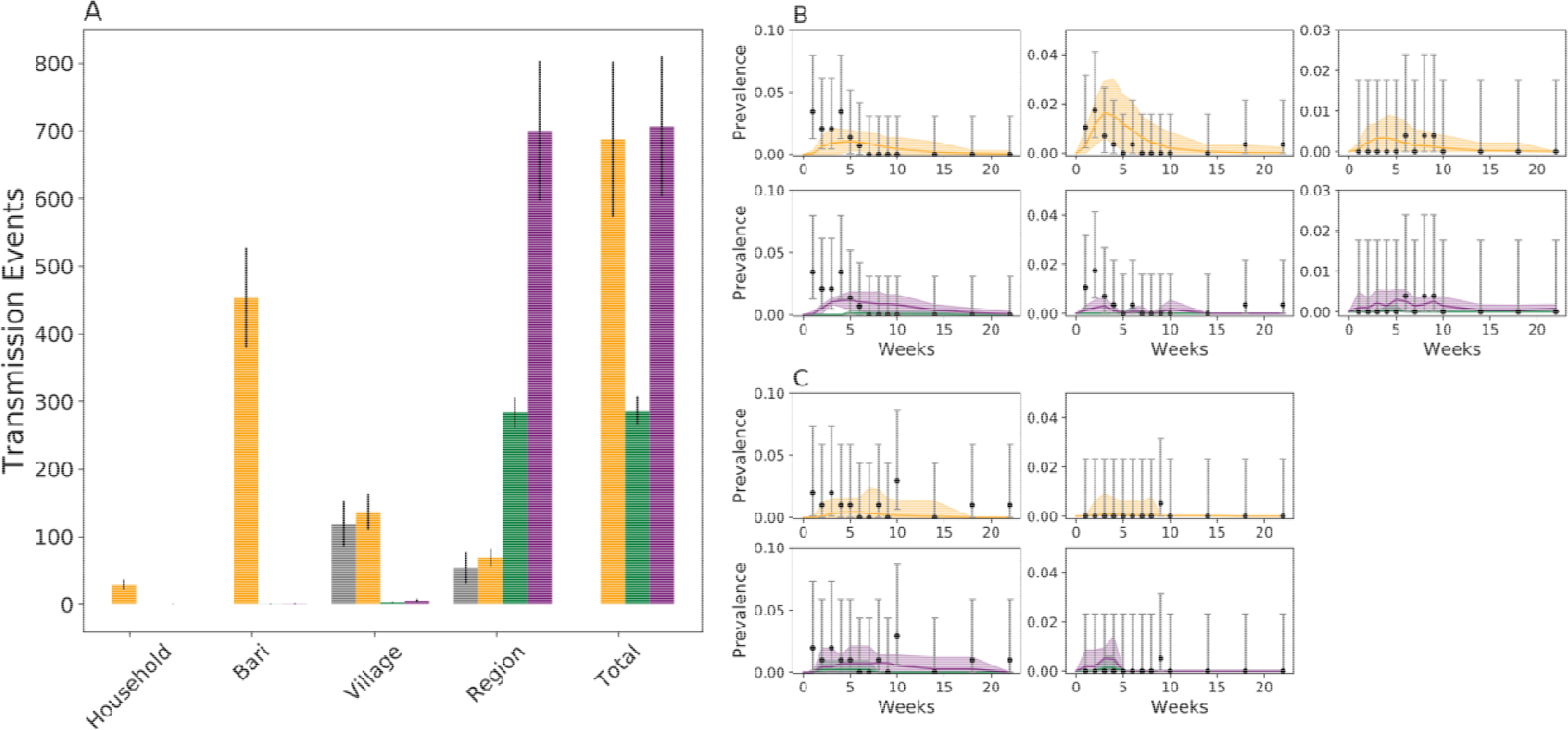
Multiscale vs Mass action calibration. *Orange*: multiscale, *Green*: fully calibration mass action, and *Purple*: partially calibrated mass action. The fully calibrated mass action model was fit to both phases of the clinical trial. The partially calibrated mass action model was fit only to the longitudinal shedding profiles collected during the post-mOPV2 challenge surveillance period. **A**) Distribution of simulated transmission events across demographic scales compared against the priors for village and regional (intervillage) transmission obtained during the routine immunization and enrollment phase (*grey*). Household and bari transmission could not be estimated form the routine immunization and enrollment phase data. Error bars indicate one standard deviation. Cohort-specific longitudinal shedding profiles due to bari/household transmission (**B**, left to right: cohorts two, six, five) or village/region transmission (**C**, left to right: cohorts three, eight). Error bars indicate two binomial standard errors. **B** and **C** are split into two rows, one for the multiscale and one for the two mass action models for visual clarity. The solid line is the simulated average and the shading indicates two standard deviations from the mean obtained from 200 simulation replicates.

Compared to the fully-calibrated mass action model, the multiscale model allocated transmissions more flexibly across demographic sublevels. Household and bari transmission comprise 0.042 (0.023, 0.054) and 0.659 (0.625, 0.693) of all simulated transmission events respectively (**Fig. 2A**, *orange*). The multiscale model predicted two times as many total transmission events as the fully-calibrated mass action model. Comparing the two model fits, multiscale transmission model better captures the clinical trial data (Akaike information criterion (AIC), 2360.14 vs 2363.08), particularly within the first ten weeks after mOPV2 challenge.

Given the failure of the fully-calibrated mass action model to replicate the clinical trial data, we also fit a mass action model to just the cohort-specific longitudinal shedding profiles collected during the post-mOPV2 surveillance period (**Fig. 2**, *purple*). The partially-calibrated mass action model represents a best-case scenario where mass action is forced to fit the clinical trial data and meant to show the consequences of differences in social contact structure. Like the fully-calibrated mass action model, most transmission events occur between villages (**Fig. 2A**) but the total number of transmission events was not significantly different form the calibrated multiscale model (706.58 vs 688.1, p-value = 0.51, Two Sample Kolmogorov-Smirnov Test, n = 200). The partially-calibrated mass action model generates better cohort-specific longitudinal shedding profiles than the fully-calibrated mass action model (AIC 2337.86 vs 2354.81) and multiscale model (AIC 2337.86 vs 2358.96) (**Fig. 2B-C**, *purple*). However, it underestimates shedding in cohort six, which is composed of non-mOPV2 challenged household contacts in close contact with a mOPV2-challenged infant (**Fig. 2B**, *center*).

### Forecasting transmission risk

Sabin 2 transmission and cVDPV2 introduction risk following Sabin 2 vaccination cessation was modeled using both the multiscale and partially-calibrated mass action models following three scenarios: 1) a single Sabin 2 importation, 2) a mOPV2 vaccination campaign with up to 40% coverage in children under five years of age, and 3) a single cVDPV2 importation. Newly born individuals in these simulations are immunologically naïve and existing immunity wanes with time since vaccination cessation (**Fig. S4**). cVDPV2 importation risk is modeled using a Type 2 WPV strain, which is characterized by longer shedding durations and higher infectivity (21). All scenarios were simulated using the multiscale and partially-calibrated mass action models.

Simulated Sabin 2 importations exhibited significant simulation to simulation variation. Epidemics could be grouped into two categories based on persistence times (**Fig. 3A**): 1) small unstable epidemics that fade out within a year, and 2) metastable epidemics that persist for longer than a year before fading out. As the time since vaccination cessation increased, metastable epidemics became more frequent. However, they were less frequent in the multiscale model, occurring in 3.6% and 15.5% of simulated point importations five- and 40-years post-vaccination cessation. The corresponding rates in the partially-calibrated mass action model were 22.5% and 30.1% respectively. The epidemics predicted by the partially-calibrated mass action model infected more individuals (**Fig. 3B-C**) than those in the multiscale model. After one year of vaccination cessation, the partially-calibrated mass action model predicted peak prevalence levels that were an order of magnitude higher than those under the multiscale model (**Fig. S5**).

**Fig. 3.**
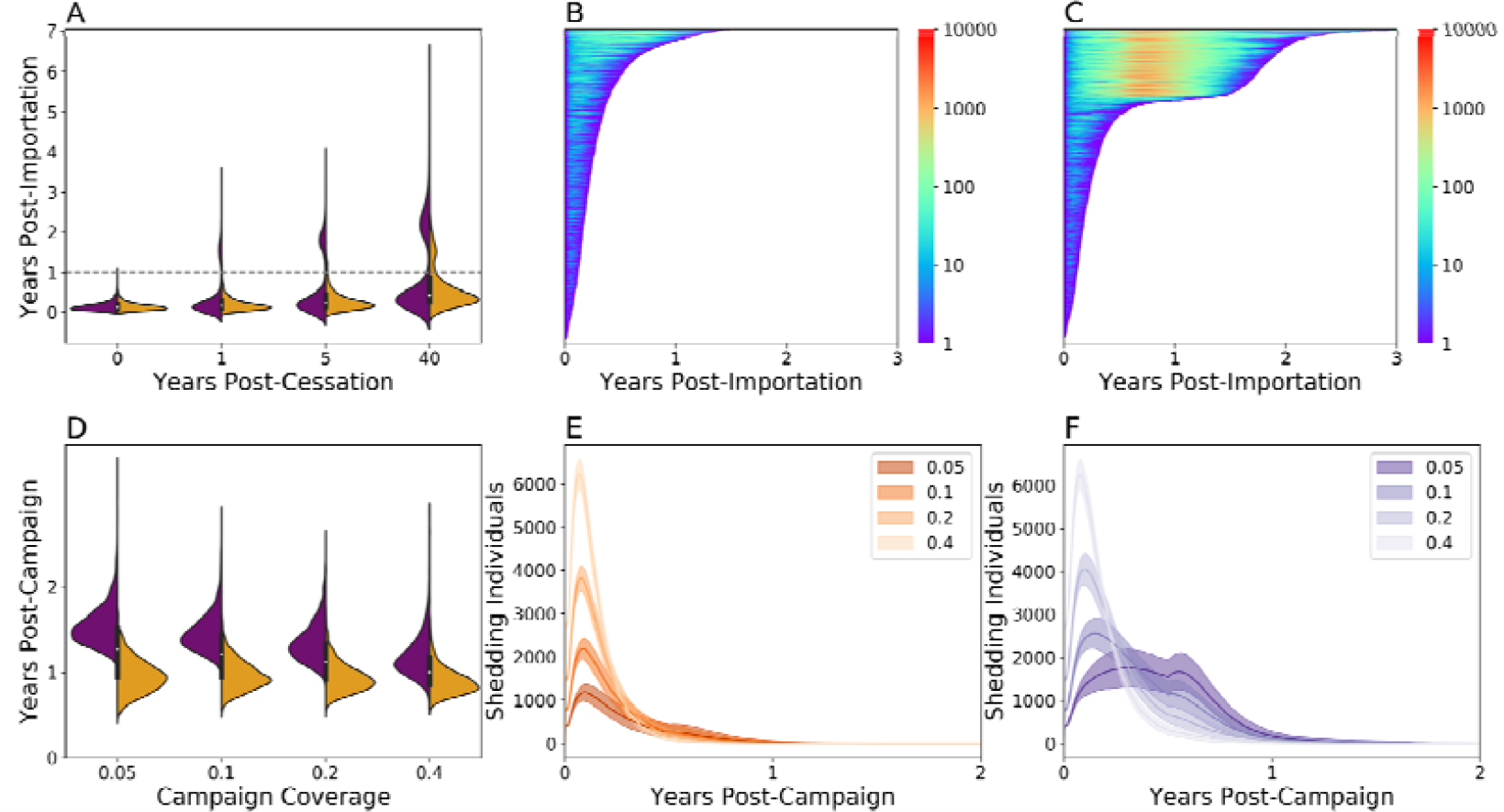
Model comparison and extrapolation. Model predictions using the multiscale (*orange*) and partially-calibrated mass action (*purple*) models for Sabin 2 point-importation outbreaks (**A-C**) or mOPV2 campaign (**D-F**). Simulations were repeated 2000 times. **A**) Transmission persistence after vaccination cessation. Individual simulation outcome following Sabin 2 point-importation outbreaks five years post-vaccination cessation using the multiscale (**B**) and partially-calibrated mass action model (**C**). Each line of the lasagna plots represents a different simulation outcome. The length of each line indicates the persistence time and the color the number of shedding individuals at that timepoint. **D**) Transmission persistence following mOPV2 campaigns with up to 40% coverage in children under five, five years post-vaccination cessation. Number of shedding individuals in these mOPV2 campaigns using multiscale transmission (**E**) and the partially-calibrated mass action model (**F**) across different campaign coverages. clarity. The solid lines are the simulated average and the shading indicates two standard deviations from the mean obtained from 200 simulation replicates.

Simulated mOPV2 vaccination campaigns did not have bimodal persistence times (**Fig. 3D**). Higher coverage mOPV2 campaigns were associated with shorter epidemics, but all epidemics seeded by these campaigns faded out, even when only 5% of children under five were vaccinated. The partially-calibrated mass action model predicted epidemics that were longer and larger than those with the multiscale model. Interestingly, the epidemiology curves from these simulations were bimodal (**Fig. 3E-F**), with an initial peak in infected individuals at 50-100 days of transmission and a second peak centered at 200-300 days of transmission. Campaigns with higher vaccination coverage had higher initial peaks and lower secondary peaks than those with lower vaccination coverage. However, the secondary peaks in the multiscale model were smaller than those in the partially-calibrated mass action model.

cVPDV2 importation were characterized by cyclical epidemiology curves that resulted in either fadeout or the establishment of endemic transmission (**Fig. 4, Fig. S6**). Endemic transmission is defined as having achieved a stable, steady-state number of infections after ten years of transmission. Under both the multiscale and partially-calibrated mass action model, endemic transmission became more frequent as the time since vaccination cessation increased. The multiscale model predicted that 3.1% and 34.9% of cVDPV2 importations resulted in endemic transmission immediately and ten years after vaccination cessation. The partially-calibrated mass action model predicted higher frequencies of endemic epidemics and shorter fadeout persistence times. It also predicted that 30.5% and 85.9% of cVDPV2 importations resulted in endemic epidemics immediately and ten years after vaccination cessation.

**Fig. 4.**
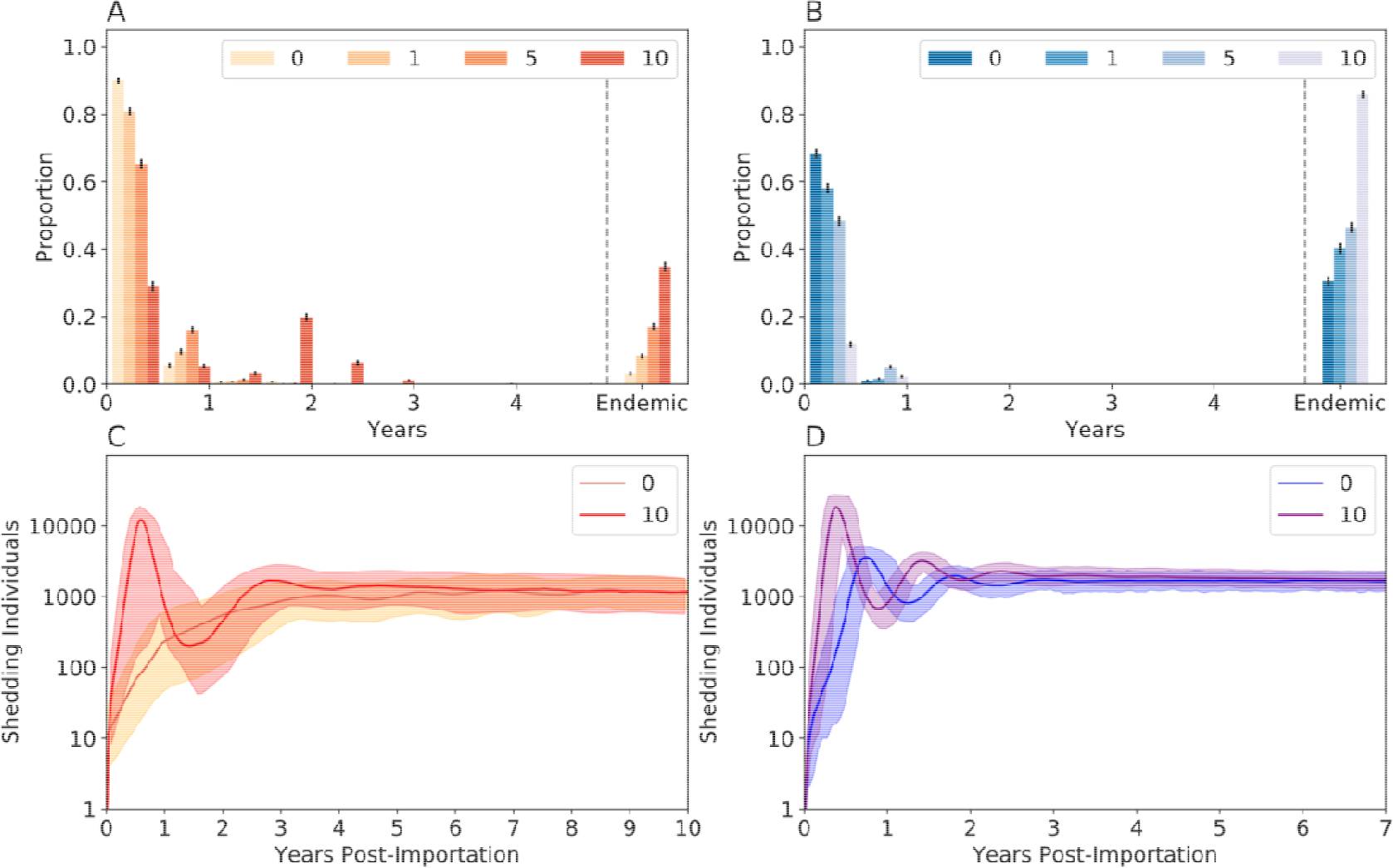
WPV Importation. Transmission persistence following WPV importation up to ten years post-vaccination cessation using the **A**) multiscale model or **B**) the partially-calibrated mass action model. Endemic transmission is defined as having any transmission for at least ten years. Error bars indicate one standard deviation. Number of shedding individuals over time for the **C**) multiscale model and **D**) partially calibrated mass action model. Only WPV importation immediately after vaccination cessation and ten years post-vaccination cessation are shown. The solid lines are the simulated average and the shading indicates two standard deviations from the mean obtained from 200 simulation replicates.

## Discussion

Accurate disease forecasting hinges on our ability to model the relationships between social contact structure and transmission (22–24). Household community structure in our multiscale model alters transmission by redistributing contacts among household, bari, village, and non-village members. This redistribution results in increased stochastic transmission failure that slows epidemic progression. When little is known about contact structure or demography, it is appealing to utilize simple models that capture some but not all features of the data. Such “partially-calibrated” models should be avoided for disease forecasting because they do not accurately capture stochastic transmission loss and will overestimate transmission and outbreak risk (16, 25, 26).

The multiscale model indicates that Sabin 2 transmission is maintained within local communities that occasionally escape to infect other villages or regions. Simulated transmissions under the multiscale model occurred predominantly between households in the same *bari* or village. While household transmission is observed following vaccination (20, 27, 28), our simulations show that they constitute a small fraction of the total transmission events because the limited number of individuals per household places an upper limit on household transmission. Sabin 2 vaccine transmission exists as series transmission events alternating between local household clusters that occasionally escape to infect more distant households throughout the region. This complex transmission structure is impossible to replicate using mass action because it incorrectly simulates all transmission as occurring between villages.

The increased stochastic transmission loss is particularly relevant when the initial number of infections is small, such as during a point importation event. These epidemics are especially prone to stochastic transmission failure and will often fail to reach the critical outbreak threshold needed to establish larger epidemics (29, 30). This was reflected by the limited emergence of metastable epidemics in the multiscale model as compared to the partially-calibrated mass action model. Interestingly, the constraints imposed by household community structure are strong enough to limit metastable epidemic emergence after forty years of vaccination cessation. Household community structure plays a critical role in managing transmission by protecting some immunologically naïve individuals from being infected with either Sabin 2 vaccine virus or cVDPV2 viral strains. Household community structure limits cVDPV2 transmission and reduces the seeding probability of cVDPV2 to new communities (12, 31). However, it also impedes the establishment of herd immunity by reducing transmission of unreverted vaccine virus from vaccinated individuals.

Household community structure also influences transmission when the initial number of shedding individuals is large, such as following a vaccination campaign. Household community structure prevents transmission from resurging after the initial wave of infections, despite the availability of immunologically naïve individuals from new births and waning immunity. Our simulations show that Matlab is not conducive to sustained Sabin 2 vaccine transmission, which is unsurprising because it is a highly immunized setting with adequate sanitation measures maintained by the installation and use of clean water tubewells and latrines (32). Despite this, Sabin 2 transmission following a mass vaccination campaign can be sustained for years, even after the additional stochastic transmission loss from household community structure is accounted for. This long transmission period increases the probability of Sabin 2 genetic reversion to cVDPV2 and highlights the importance of preventing exportation to nearby regions. This long-term residual transmission is likely a contributing factor to how mOPV2 campaigns can be responsible for over half of the 41 cVDPV2 outbreaks observed since 2016 (2).

For cVDPV2, the constraints imposed by household and community membership are counteracted by a higher R_0_ resulting from higher infectivity and longer shedding durations (21). We show that Matlab has a non-zero risk of establishing sustained cVDPV2 transmission following importation with a cVDPV2 virus. As global immunity against Sabin 2 declines, cVDPV2 importations will have higher probabilities of establishing endemic transmission. Transient cVDPV2 outbreaks will also become more frequent and last for longer periods of time before fading out. Exportation of cVDPV2 from outbreak regions must be prevented. We estimate that Matlab currently has a 20% chance of establishing endemic cVDPV2 that will double in the next five years. Regions with poorer sanitation measures will have even higher risks.

As immunity against type 2 poliovirus declines, controlling and preventing cVDPV2 outbreaks will become increasingly important, especially as the current tools and resources for combating cVDPV2 outbreaks become limited. Disease forecasting models are invaluable tools for assessing future risk, but must incorporate biological realism regarding host demographic structure, immunity, transmissibility to ensure the effectiveness of future vaccination campaigns or public health interventions. To fully assess cVDPV2 emergence risk, future iterations will need to simulate Sabin 2 genetic reversion, the use of the novel oral polio vaccines (33), and the incorporation of other social factors that could improve model fit (24). Our model establishes a framework for simulating transmission in rural communities defined by household community structure and can be reparametrized to study diseases including cholera, typhoid, and shigellosis in other study populations.

## Materials and Methods

### Study Trial Data

Stool samples were collected during both the routine immunization and enrollment phase as well as the post mOPV2 surveillance period (20). Stool samples collected during the routine immunization and enrollment phase from both tOPV- and bOPV-assigned villages were used to obtain priors for intervillage and within-village transmission rates (**SI Appendix: Calibrating Transmission**). Stool samples collected during the post-mOPV2 surveillance period from bOPV-assigned villages were used to construct eight cohort-specific, longitudinal shedding profiles designed to identify within-household, within-bari, within-village, and intervillage transmission (**Table S1**). These samples were collected at weekly intervals for the first ten weeks following the mOPV2 campaign and at weeks 14, 18, and 22 thereafter. The post-mOPV2 surveillance period stool samples from tOPV-assigned villages were not included because positive stool samples are more likely to have originated from routine immunization than those collected from bOPV-assigned villages. There were no spatial patterns in study enrollment or routine immunization assignment.

This clinical trial was done according to the guidelines of the Declaration of Helsinki. The protocol was approved by the Research Review Committee (RRC) and Ethical Review Committee (ERC) of the icddr,b and the Institutional Review Board of the University of Virginia.

### Model Overview

Our model consists of three models: 1) a poliovirus dose-response model, 2) a household demographic model, and 3) a contact-based transmission model (**Fig. 1**). The poliovirus dose-response model summarizes the existing knowledge of within-host immunity, shedding and susceptibility for each of the Sabin vaccine strains and WPV (21) (**SI Appendix: Modeling Immunity**). The shedding duration and infectivity of these viruses are also described by this dose-response model (21). Both shedding duration and the infectivity depends on host immunity. Sabin 2 has a shorter shedding duration and lower infectivity than WPV (21). This model simulates individual susceptibility to infection and transmission risk using a mathematical construct of protective, humoral immunity (the OPV-equivalent antibody titer). Infected individuals with high OPV-equivalent antibody titers are less susceptible to infection, have shorter infection durations, and shed less virus per gram of stool than those with smaller OPV-equivalent antibody titers.

The household demographic model is based on an anthropological framework of household formation and secession (34). Individuals are grouped into three nested hierarchies: 1) households, 2) *baris*, and 3) villages, which together make up the Matlab study population. Households evolve through birth death processes that governs its composition over time. Individuals can move between households through marriage or leave to start their own households. These dynamics are governed by fertility (age specific fertility rate, birth interval period, children per married female, and child preference number), mortality, and marriage rates obtained from the demographic data reported by the WHO (35) and the Bangladesh Demographic Health Surveys (BDHSS) (36, 37) (**SI Appendix: Modeling individuals, SI Appendix: Household demographic structure**). Rules regarding household secession and formation were informed by reports of rural Bangladeshi society (38) and calibrated to the household size distributions reported in the BDHSS.

When simulating transmission, infected individuals contact and expose others to infectious virus. Successful transmission depends on the viral shedding concentration of the transmitter and the susceptibility of the recipient. Infectious contacts are allocated using two contact frameworks. The first assumes mass action and distributes contacts randomly throughout the entire population using a single daily contact rate, β_ma_. The second assumes multiscale transmission. Infected individuals randomly contact household members, bari members, village members, and non-village members. The number of contacts per demographic scale per day was defined using four independent contact rates (β_household_, β_bari_, β_village_, and β_intervillage_). These contact frameworks were each independently calibrated to the empirical estimates of within-village and intervillage transmission based on stool samples collected during routine immunization and enrollment as well as the cohort-specific, longitudinal shedding profiles collected after the mOPV2 campaign **SI Appendix: Calibrating Transmission**).

### Clinical Trial Simulation

The conditions of the mOPV2 clinical trial were replicated by:

1. Assigning immunity to infants from bOPV routine immunization and non-infant immunity assuming repeated, non-specific immunization source representing an unknown mix of vaccination, vaccine virus transmission, and wild poliovirus activity (20, 39) (**SI Appendix: Modeling Immunity**). Secondary immunization from routine immunization was not simulated.
2. Randomly enrolling half the infants in each village to be monitored for Sabin 2 shedding. A third of the enrolled infants received mOPV2.
3. Enrolling the two household contacts of each enrolled infant. Priority was given to the two youngest individuals < 14 years of age. If not possible, the next youngest female household members were chosen until two household contacts could be obtained, which reflects the sampling used in the study (20). Five percent of enrolled household contacts were challenged with mOPV2.
4. Challenging 40% of all children < five years of age with mOPV2
5. Simulating onward transmission assuming mass action or multiscale transmission in single day time steps for 22 weeks. Changes in household community structure are also updated.

Vaccination cessation was simulated by assuming no vaccination or transmission for a period of up to 40 years. During this period, fertility and mortality rates were assumed to be the same as those in 2014.

## Data Availability

Data requests regarding the clinical trial should be made to the Dr. Mami Taniuchi. This trial is registered at ClinicalTrials.gov, number NCT02477046. The model and code used in this study will be available on Github (https://github.com/InstituteforDiseaseModeling/MultiscaleModeling.git) at the time of publication.

## Acknowledgments

We acknowledge the International Centre for Diarrhoeal Disease Research and the Matlab community for their support and consent. We also thank John F. Modlin at the Bill and Melinda Gates Foundation for manuscript comments and suggestions. This study was funded by the Bill & Melinda Gates Foundation (OPP112545 to Petri and Taniuchi, OPP1198444 to Taniuchi).

